# Autofluorescence of the parathyroid glands during total thyroidectomy. Review of systematic reviews

**DOI:** 10.1101/2024.12.21.24319459

**Authors:** José Luis Pardal-Refoyo, Beatriz Pardal-Pelaéz

## Abstract

**Introduction and Objective:** This study reviews the use of near-infrared autofluorescence (NIRAF) for identifying parathyroid glands during total thyroidectomy. The main goal is to evaluate whether using NIRAF reduces the incidence of postoperative hypocalcemia, a common complication of thyroid surgery. The study analyzes previously published meta-analyses to assess the effectiveness of NIRAF compared to traditional visual identification methods. The preservation of parathyroid glands during thyroid surgery is essential to prevent hypoparathyroidism and consequent hypocalcemia.

**Methods:** The research involved a review of systematic reviews and meta-analyses found in PubMed, WoS, and the Cochrane Library databases. Three meta-analyses that included studies on total thyroidectomy with and without NIRAF were selected. These meta-analyses evaluated transient and permanent postoperative hypocalcemia. The data from 15 studies included in the meta-analyses were extracted and statistically reanalyzed (meta-meta-analysis). The statistical analysis was performed using the ESCI statistical package of the JAMOVI program and METAFOR by JASP. The PRISMA guidelines were followed.

**Results:** The use of NIRAF during total thyroidectomy was associated with a significantly lower incidence of both global and transient hypocalcemia. The prevalence of permanent hypocalcemia was also lower in the NIRAF group, but the difference was not statistically significant. The meta-meta-analysis showed a statistically significant association between the use of NIRAF and a reduction in hypocalcemia. There was moderate to high heterogeneity in errors and possible asymmetry in the data for global and transient hypocalcemia, but not for permanent hypocalcemia.

**Conclusions:** NIRAF is a useful tool for identifying and preserving parathyroid glands during total thyroidectomy, and its use is associated with a reduced risk of postoperative hypocalcemia. The current evidence supports the use of NIRAF, although the strength of the recommendation is weak because of heterogeneity, risk of bias, and inconsistent results among the studies. More well-designed studies are needed to confirm these findings and establish NIRAF as a standard.

## Introduction

The identification, preservation of your vascularization, and preservation of the parathyroid (PG) glands during thyroid and parathyroid surgery aims to prevent hypoparathyroidism and consequent postoperative hypocalcemia with significant consequences for the patient’s health [1–3].

Usually the localization of PGs in thyroidectomy depends on visual identification related to the surgeon’s experience and the recognition of subtle anatomical features, resulting in a considerable rate of damaged or accidentally removed PGs, especially in the hands of less experienced surgeons [2].

The search for a more accurate and reliable technique for intraoperative identification of PGs has led to the development of near-infrared autofluorescence (NIRAF) which offers significant advantages over traditional methods of identifying PGs [2–4].

NIRAF is based on the intrinsic ability of PGs to emit a fluorescent signal when illuminated with near-infrared light, allowing PGs to be visualized in real time, facilitating their identification and preservation during surgery even before they are recognizable to the naked eye, minimizing the risk of damage during manipulation of surrounding tissues, especially in cases of distorted anatomy, such as in bulky goiters or in reoperations [1,2].

Autofluorescence is a physical phenomenon in which molecules called fluorophores emit light after being excited by electromagnetic radiation, such as ultraviolet or visible light. This process occurs when a fluorophore absorbs a photon, raising its energy level to an excited state. Subsequently, the fluorophore returns to its ground state, part of the energy is dissipated in the form of heat, releasing the absorbed energy in the form of a photon of lower energy, that is, of a longer wavelength with a very short lifetime of the excited state, in the order of nanoseconds, and the intensity of the fluorescence depends on the concentration of the fluorophore and the efficiency of the process [2].

The parathyroid glands exhibit strong autofluorescence under near-infrared (NIR) light significantly greater than that of surrounding tissues, such as the thyroid gland and adipose tissue [1,2] which has allowed the development of techniques for the identification of these glands during thyroid and parathyroid surgery. Devices such as Fluobeam® 800, which uses a NIR laser with a wavelength of 750 nm and filters that select wavelengths between 800 and 850 nm, corresponding to the autofluorescence of parathyroids [4].

The molecular origin of autofluorescence in the parathyroid glands is still unknown [2]. Autofluorescence has been speculated to originate from specific cellular structures, such as the extracellular calcium-sensitive receptor, but more studies are needed to confirm this hypothesis [4].

NIRAF has a high sensitivity and specificity for the detection of PGs [2,3] as demonstrated in the meta-analysis conducted by Wang et al. (2021) that confirmed the accuracy of NIRAF, consolidating its position as a reliable tool for the identification of PGs [3]. The overall combined sensitivity and specificity of NIRAF were 96%, with an area under the curve (AUC) of 0.99 and subgroup analysis revealed that the accuracy of NIRAF was influenced by the measurement instrument, the type of disease, and the reference standard used for PG diagnosis [3].

It is a non-invasive technique that does not require the administration of substances, making it a safe and simple technique to integrate into the surgical workflow, ideal for surgeon training, as it allows less experienced surgeons to become familiar with the appearance of PGs and gain confidence in their identification. improving the learning curve and promoting patient safety [2].

NIRAF can have drawbacks such as interference with other tissues with autofluorescent properties similar to those of PGs, such as brown fat or lymph nodes that can cause false positives or limited light penetration that can make it difficult to visualize PGs located in depth (false negatives) and in certain pathologies such as in patients with secondary hyperparathyroidism or in multiple endocrine neoplasia type 1[1,2].

On the other hand, NIRAF alone does not provide information on the vascularization or functionality of PG, so complementary techniques, such as indocyanine green angiography (ICG), are needed to assess its viability [1].

There are several commercially available systems that use autofluorescence for the identification of parathyroid glands. Some use a fiber-optic probe that is placed in contact with tissue to detect the autofluorescence signal, such as PTeye®. Other systems rely on a camera to capture fluorescence images of the surgical field, such as the Fluobeam 800®, Elevision® IR, and PDE Neo II.® In addition, there are systems such as NIR fluorescence glasses or the Tissue Overlay Imaging System (OTIS) [1,2].

Therefore, during total thyroidectomy, autofluorescence of the parathyroid glands has some advantages such as help in the identification of the parathyroid glands even before visualization with the naked eye, higher visualization rate in published studies, reduction of the rate of hypocalcemia, aid in decision-making and as a tool during teaching [2,4].

However, as with all technological innovations, it is difficult to change routines and make the necessary initial financial investments. Clinicians and hospital managers need information about the benefits of the new technique.

Among others, one of the improvements that should be evaluated is whether the use of NIRAF in total thyroidectomy is associated with a lower incidence of hypocalcemia in the postoperative period as an overall indicator of improvement in the quality of patient care.

The aim of this study is to review the results of published systematic reviews on the use of parathyroid gland autofluorescence during total thyroidectomy by evaluating the rate of transient and permanent post-operative hypoparathyroidism.

## Methods

Review of systematic reviews with meta-analyses on the identification of parathyroid glands with autofluorescence during total thyroidectomy in the PubMed (https://pubmed.ncbi.nlm.nih.gov/), WoS (https://www.webofscience.com/wos/woscc/basic-search) and Cochrane Library (https://www.cochranelibrary.com/) databases with search *strategies((autofluorescence) AND (parathyroid glands)) DNA (total thyroidectomy)*.

Inclusion criteria: meta-analyses evaluating autofluorescence of the parathyroid glands in total thyroidectomy with data on hypocalcemia or total, transient, and permanent postoperative hypoparathyroidism.

Exclusion criteria: series with fluorescence techniques using only fluorescent substances, patients with hemithyroidectomy, two-stage total thyroidectomy (totalization), revision thyroid surgery, previous cancer treatment, and patients with secondary or recurrent cancer.

The PRISMA guidelines were followed for the study [5] (https://www.prisma-statement.org/prisma-2020).

In each of the meta-analyses, the following were analyzed: the variables evaluated, heterogeneity, publication bias, comparison of results, research biases, methodological quality, level of evidence and degree of recommendation with the GRADE system (https://book.gradepro.org/), each meta-analysis was evaluated citrically using the CASPe guidelines for systematic reviews (https://redcaspe.org/materiales/).

Subgroups in which NIRAF was used (experimental) versus those in which NIRAF was not used (control) were evaluated and in them the total number of hypocalcemia, the number of patients with transient hypocalcemia, and the number of patients with permanent hypocalcemia.

Each meta-analysis was assessed for formal quality using the checklist according to PRISMA guidelines.

A narrative synthesis and meta-meta-analysis was performed with the trials included in the meta-analyses selected with the ESCI statistical package of the JAMOVI program (The jamovi project (2024). (Version 2.6) [Computer Software], https://www.jamovi.org) and METAFOR by JASP (JASP Team (2024). JASP (Version 0.19.1) [Computer software], https://jasp-stats.org/download/) with the method of maximum likelihood with random effects.

NotebookLM (https://notebooklm.google.com, December 2024) was used for text analysis and synthesis table generation.

## Results

The tables and figures with the results can be consulted in the Annex.

The PRISMA diagram in Figure 1 summarizes the strategy for searching and selecting articles [6].

**Figure 1.**
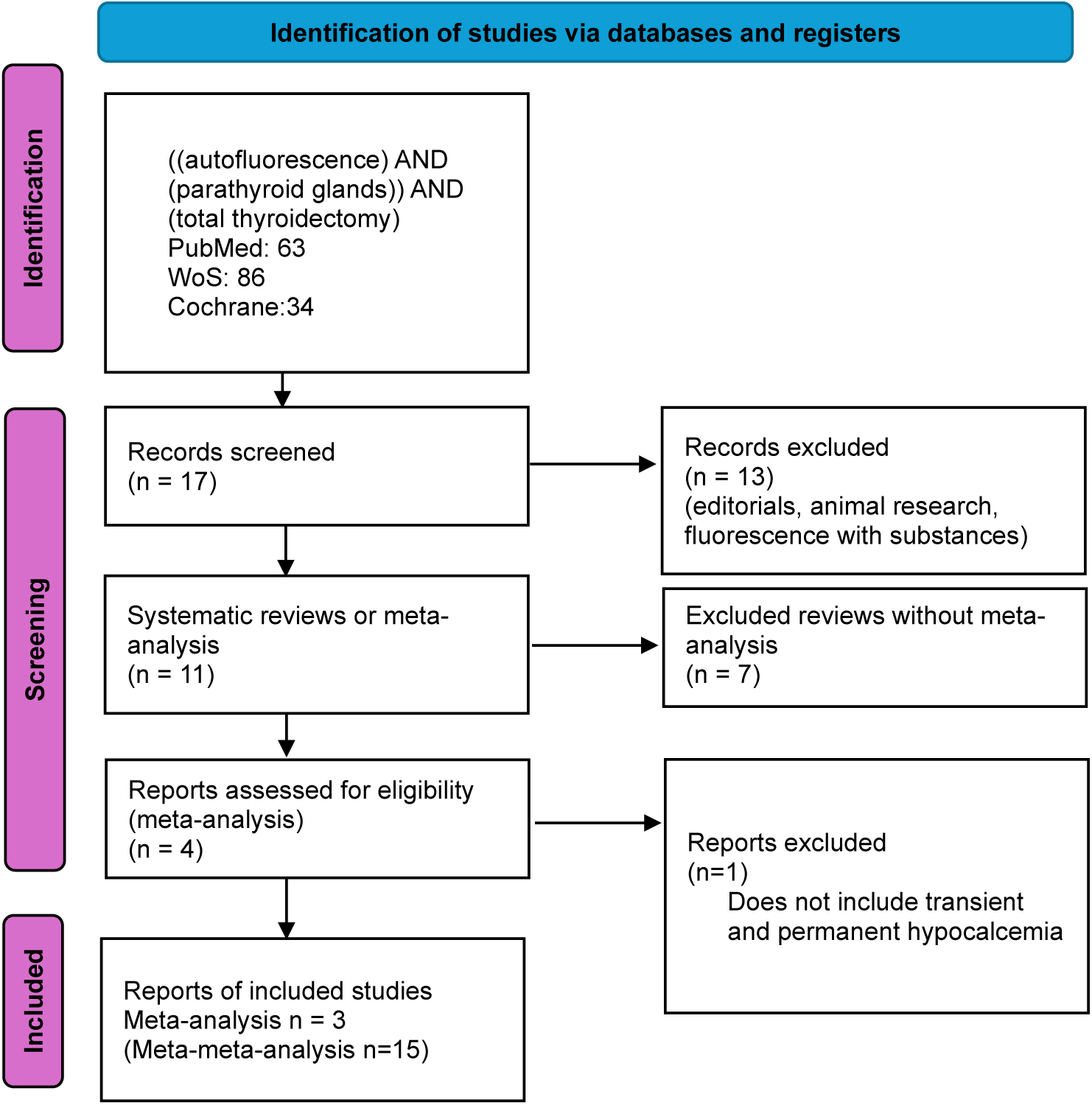
PRISMA diagram with the selection of articles.

We selected three meta-analyses that include studies on measuring the outcome of transient and permanent postoperative hypocalcemia after performing total thyroidectomy with NIRAF (NIRAF group) or without NIRAF (control group) [7–9].

The general characteristics of the meta-analyses are summarized in Table 1 and their main results in Table 2 (rate of hypocalcemia or hypoparathyroidism).

**Table 1.** Number of patients included in the meta-analyses.

| Meta-analysis | included studies | Total N | n NIRAF | n control |
| --- | --- | --- | --- | --- |
| Rao et al. (2024) [7] | RCTs [14] [10] [19] [18] [17] [15] [23] [24] [20] | 1363 | 636 | 637 |
| Safia et al. (2024) [8] | RCTs [10] [14] [18] [20] [12] [17] [23] [24] [21] | 1711 | 856 | 855 |
| Weng et al. (2021) [9] | RCTs and observational studies [11] [10] [13] [16] [22] [14] [17] | 2180 | 493 | 1687 |

**Table 2.**
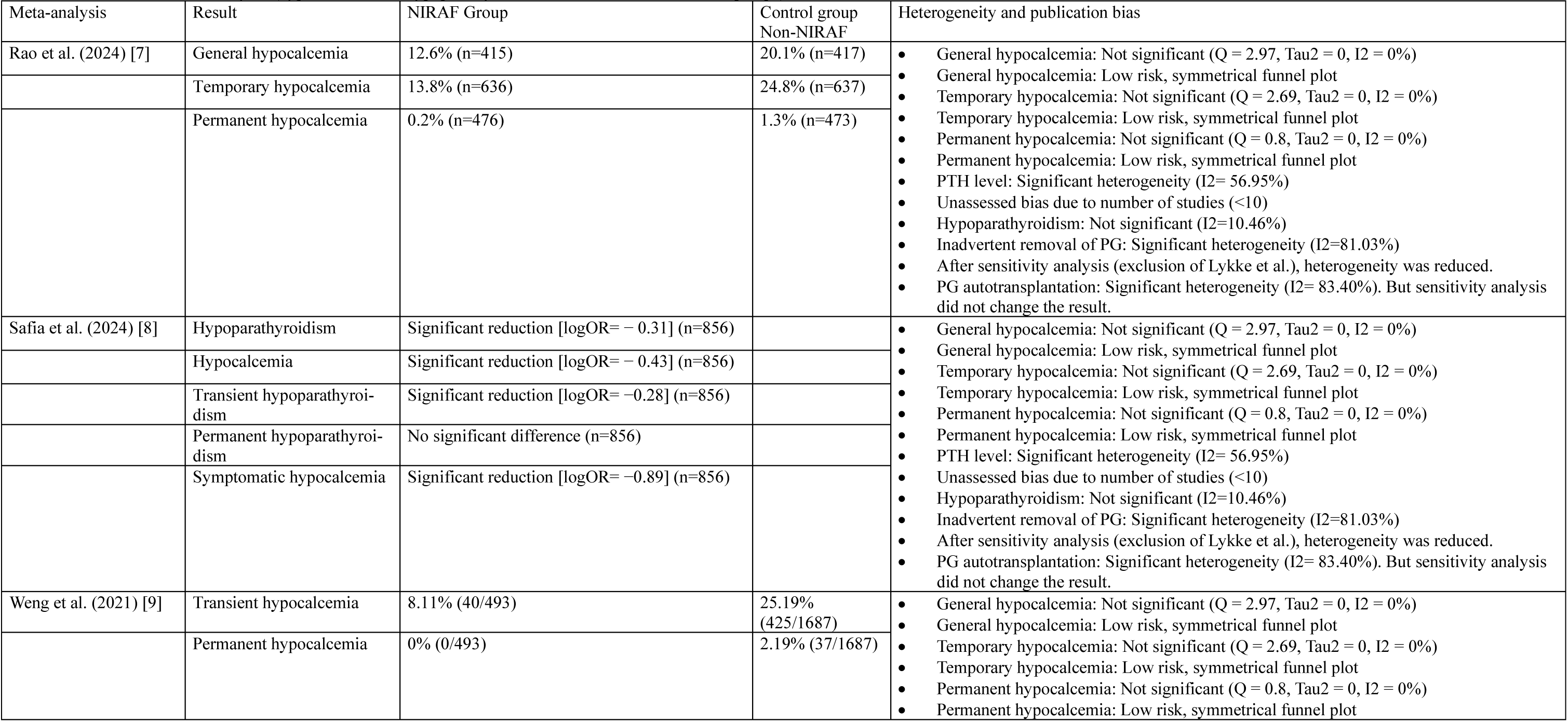
Main results of meta-analyses (hypocalcaemia or hypoparathyroidism in the NIRAF and control groups).

The selected meta-analyses include 15 studies whose results were extracted from the original documents and statistically analysed (meta-meta-analysis) [10–24]. Table 3 summarizes the general results related to the type of research, NIRAF technology used.

**Table 3.**
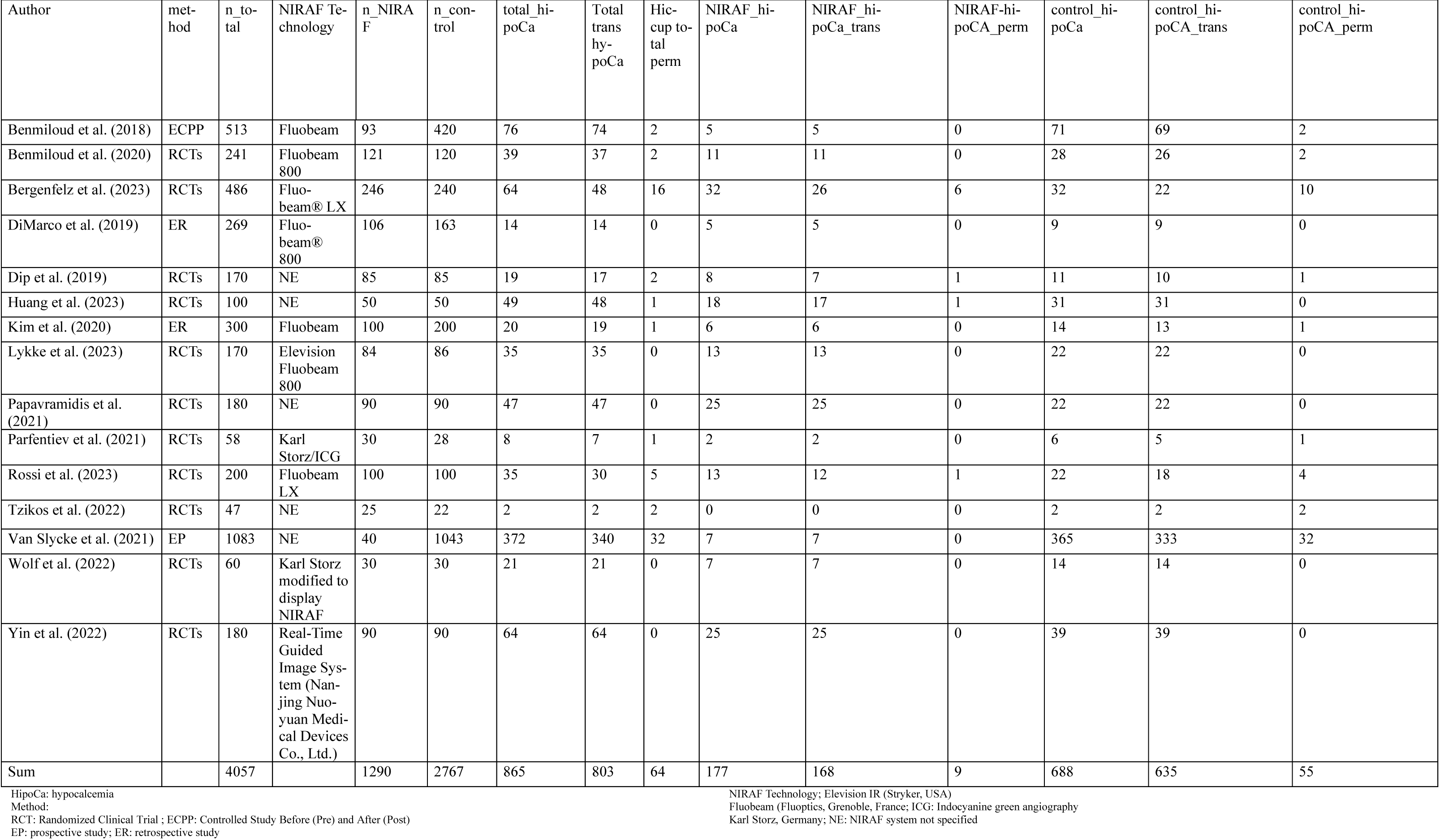
Number of patients in the 15 included studies. [10–24] **in meta-analyses** [7–9].

Table 4 shows the results of hypocalcemia (general for the sample, transient and permanent) in the NIRAF (experimental) and control (without NIRAF) groups. with the proportions and their 95% CI.

**Table 4.** Analysis of proportions. Hypocalcemia in each study (total, transient, and permanent)

|  |  | HypoCa in the sample |  |  |  | Transient hypoCa in the sample |  |  |  | Permanent HypoCa in the Specimen |  |  |  |
| --- | --- | --- | --- | --- | --- | --- | --- | --- | --- | --- | --- | --- | --- |
|  |  |  |  | 95% IQ |  |  |  | 95% IQ |  |  |  | 95% IQ |  |
| Study label | N | Cases | P | LL | UL | Cases | P | LL | UL | Cases | P | LL | UL |
| Benmiloud et al. (2018) | 513 | 76 | 0.1481 | 0.12002 | 0.1817 | 74 | 0.1442 | 0.11648 | 0.1775 | 2 | 0.0039 | 1.84E-04 | 0.0153 |
| Benmiloud et al. (2020) | 241 | 39 | 0.1618 | 0.12061 | 0.2141 | 37 | 0.1535 | 0.11337 | 0.205 | 2 | 0.0083 | 4.58E-04 | 0.0322 |
| Bergenfelz et al. (2023) | 486 | 64 | 0.1317 | 0.10447 | 0.1649 | 48 | 0.0988 | 0.07524 | 0.1288 | 16 | 0.03292 | 0.02008 | 0.0534 |
| DiMarco et al. (2019) | 269 | 14 | 0.052 | 0.03074 | 0.0865 | 14 | 0.052 | 0.03074 | 0.0865 | 0 | 0 | 0 | 0.0174 |
| Dip et al. (2019) | 170 | 19 | 0.1118 | 0.07229 | 0.1691 | 17 | 0.1 | 0.06285 | 0.1555 | 2 | 0.01176 | 7.21E-04 | 0.0453 |
| Huang et al. (2023) | 100 | 49 | 0.49 | 0.39431 | 0.5865 | 48 | 0.48 | 0.38475 | 0.5768 | 1 | 0.01 | 0 | 0.061 |
| Kim et al. (2020) | 300 | 20 | 0.0667 | 0.04324 | 0.1015 | 19 | 0.0633 | 0.04057 | 0.0976 | 1 | 0.00333 | 0 | 0.021 |
| Lykke et al. (2023) | 170 | 35 | 0.2059 | 0.15185 | 0.2734 | 35 | 0.2059 | 0.15185 | 0.2734 | 0 | 0 | 0 | 0.0273 |
| Papavramidis et al. (2021) | 180 | 47 | 0.2611 | 0.20244 | 0.3302 | 47 | 0.2611 | 0.20244 | 0.3302 | 0 | 0 | 0 | 0.0259 |
| Parfentiev et al. (2021) | 58 | 8 | 0.1379 | 0.06974 | 0.2528 | 7 | 0.1207 | 0.05748 | 0.2328 | 1 | 0.01724 | 0 | 0.1018 |
| Rossi et al. (2023) | 200 | 35 | 0.175 | 0.1285 | 0.2342 | 30 | 0.15 | 0.10696 | 0.2068 | 5 | 0.025 | 0.00933 | 0.0593 |
| Tzikos et al. (2022) | 47 | 2 | 0.0426 | 0.00465 | 0.1522 | 2 | 0.0426 | 0.00465 | 0.1522 | 2 | 0.04255 | 0.00465 | 0.1522 |
| Van Slycke et al. (2021) | 1083 | 372 | 0.3435 | 0.31582 | 0.3723 | 340 | 0.3139 | 0.28702 | 0.3422 | 32 | 0.02955 | 0.02093 | 0.0416 |
| Wolf et al. (2022) | 60 | 21 | 0.35 | 0.24182 | 0.4769 | 21 | 0.35 | 0.24182 | 0.4769 | 0 | 0 | 0 | 0.0739 |
| Yin et al. (2022) | 180 | 64 | 0.3556 | 0.2894 | 0.428 | 64 | 0.3556 | 0.2894 | 0.428 | 0 | 0 | 0 | 0.0259 |
| **Total** | **4057** | **865** | **0.198** | **0.139** | **0.257** | **803** | **0.188** | **0.132** | **0.243** | **64** | **0.00974** | **0.00289** | **0.0166** |

HypoCa: hypocalcemia

Table 5 summarizes the number of patients included in the studies in relation to postoperative hypocalcemia. The Chi2 analysis indicates that there is a statistical association between the NIRAF and control groups in terms of the prevalence of global, transient and permanent hypocalcemia.

**Table 5.** Total number of patients enrolled in the studies [10–24]. Chi2 analysis.

|  | NIRAF | without NIRAF<br>(control) | Total | chi2 |  |
| --- | --- | --- | --- | --- | --- |
| n total | 1290 | 2767 | 4057 |  |  |
| Hiccups | 177 | 688 | 865 | 64.47 | P: 9.82E-16 |
| Transient hypoCa | 168 | 635 | 803 | 53.98 | P: 2.03E-13 |
| Permanent hypoCa | 9 | 55 | 64 | 8.62 | p: 0.0033 |
| without hiccups | 1113 | 2079 | 3192 |  |  |

HipoCa: hypocalcemia

Table 6 summarizes the prevalence of total, transient, and permanent hypocalcemia in the sample and in the NIRAF and control groups (without NIRAF) by means of proportional analysis. In the total sample, the incidence of hypocalcemia was 19.8% (95% CI: 13.9%), in the NIRAF group it was 5.28% (95% CI: 3.52% - 7.03%) and in the control group it was 13.5% (95% CI: 8.38% - 18.7%). In the total sample, the incidence of transient hypocalcemia was 18.8% (95% CI: 13.2% - 24.3%), while in the NIRAF group it was 5.0% (95% CI: 3.32% - 6.68%), and in the control group it was 12.8% (95% CI: 7.93% - 17.6%). In the total sample, the incidence of permanent hypocalcemia was 0.974% (95% CI: 0.289% - 1.66%), while in the NIRAF group it was 0.0571% (95% CI: −0.144% - 0.258%), and in the control group it was 0.8% (95% CI: 0.196% - 1.4%).

**Table 6.**
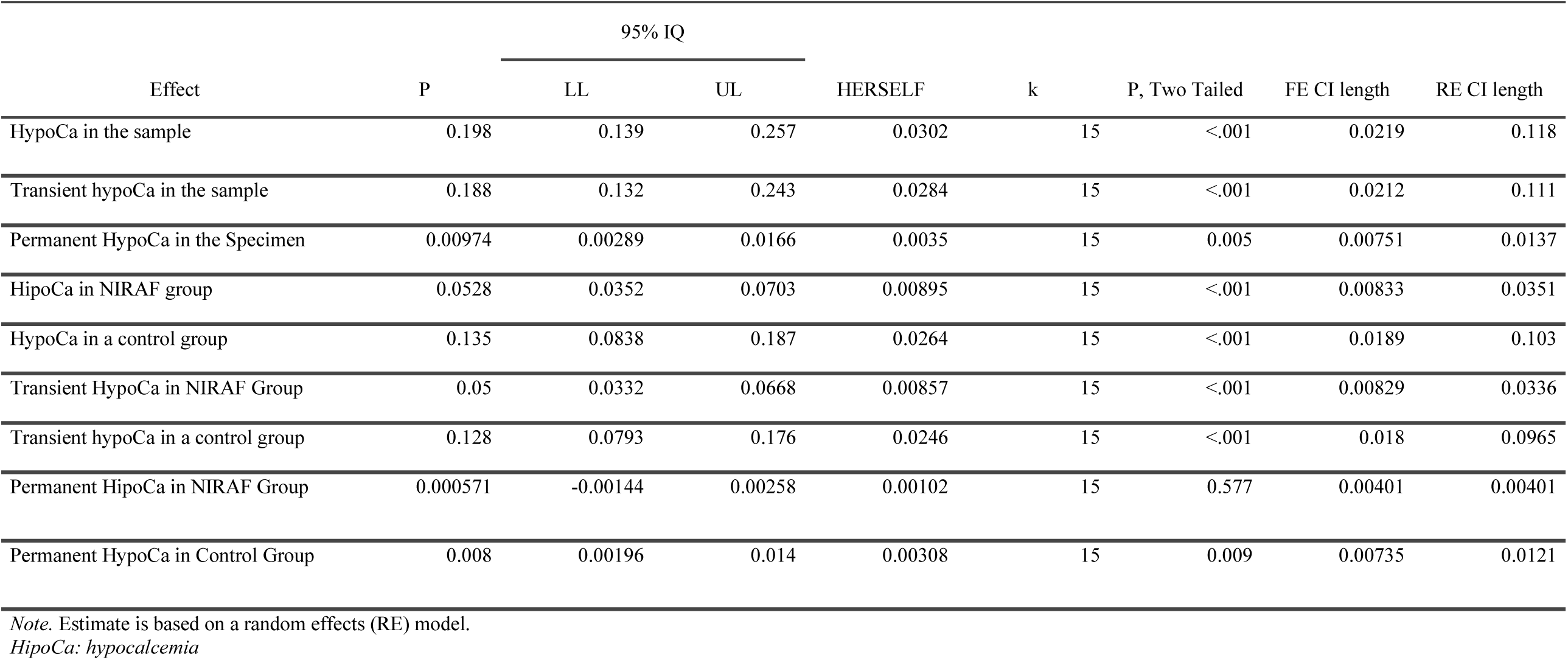
Analysis of proportions. Prevalence of total, transient and permanent hypocalcemia in the sample and in the NIRAF and control groups (without NIRAF) in the 15 selected studies.

| Effect | P | 95% IQ |  | HERSELF | k | P, Two Tailed | FE CI length | RE CI length |
| --- | --- | --- | --- | --- | --- | --- | --- | --- |
|  |  | LL | UL |  |  |  |  |  |
| HypoCa in the sample | 0.198 | 0.139 | 0.257 | 0.0302 | 15 | <.001 | 0.0219 | 0.118 |
| Transient hypoCa in the sample | 0.188 | 0.132 | 0.243 | 0.0284 | 15 | <.001 | 0.0212 | 0.111 |
| Permanent HypoCa in the Specimen | 0.00974 | 0.00289 | 0.0166 | 0.0035 | 15 | 0.005 | 0.00751 | 0.0137 |
| HipoCa in NIRAF group | 0.0528 | 0.0352 | 0.0703 | 0.00895 | 15 | <.001 | 0.00833 | 0.0351 |
| HypoCa in a control group | 0.135 | 0.0838 | 0.187 | 0.0264 | 15 | <.001 | 0.0189 | 0.103 |
| Transient HypoCa in NIRAF Group | 0.05 | 0.0332 | 0.0668 | 0.00857 | 15 | <.001 | 0.00829 | 0.0336 |
| Transient hypoCa in a control group | 0.128 | 0.0793 | 0.176 | 0.0246 | 15 | <.001 | 0.018 | 0.0965 |
| Permanent HipoCa in NIRAF Group | 0.000571 | -0.00144 | 0.00258 | 0.00102 | 15 | 0.577 | 0.00401 | 0.00401 |
| Permanent HypoCa in Control Group | 0.008 | 0.00196 | 0.014 | 0.00308 | 15 | 0.009 | 0.00735 | 0.0121 |

*Note.* Estimate is based on a random effects (RE) model.
*HipoCa: hypocalcemia*

Meta-analyses of the results of the 15 selected articles were performed to compare the differences in proportions between the NIRAF and control groups in terms of total hypocalcemia, transient hypocalcemia, and permanent hypocalcemia.

The results of the hypocalcemia analysis in the sample using the difference in proportions between the NIRAF group and the control group are summarized in Table 7, Figure 2 (Forest plot) and Figure 3 (Funnel plot).

**Figure 2.**
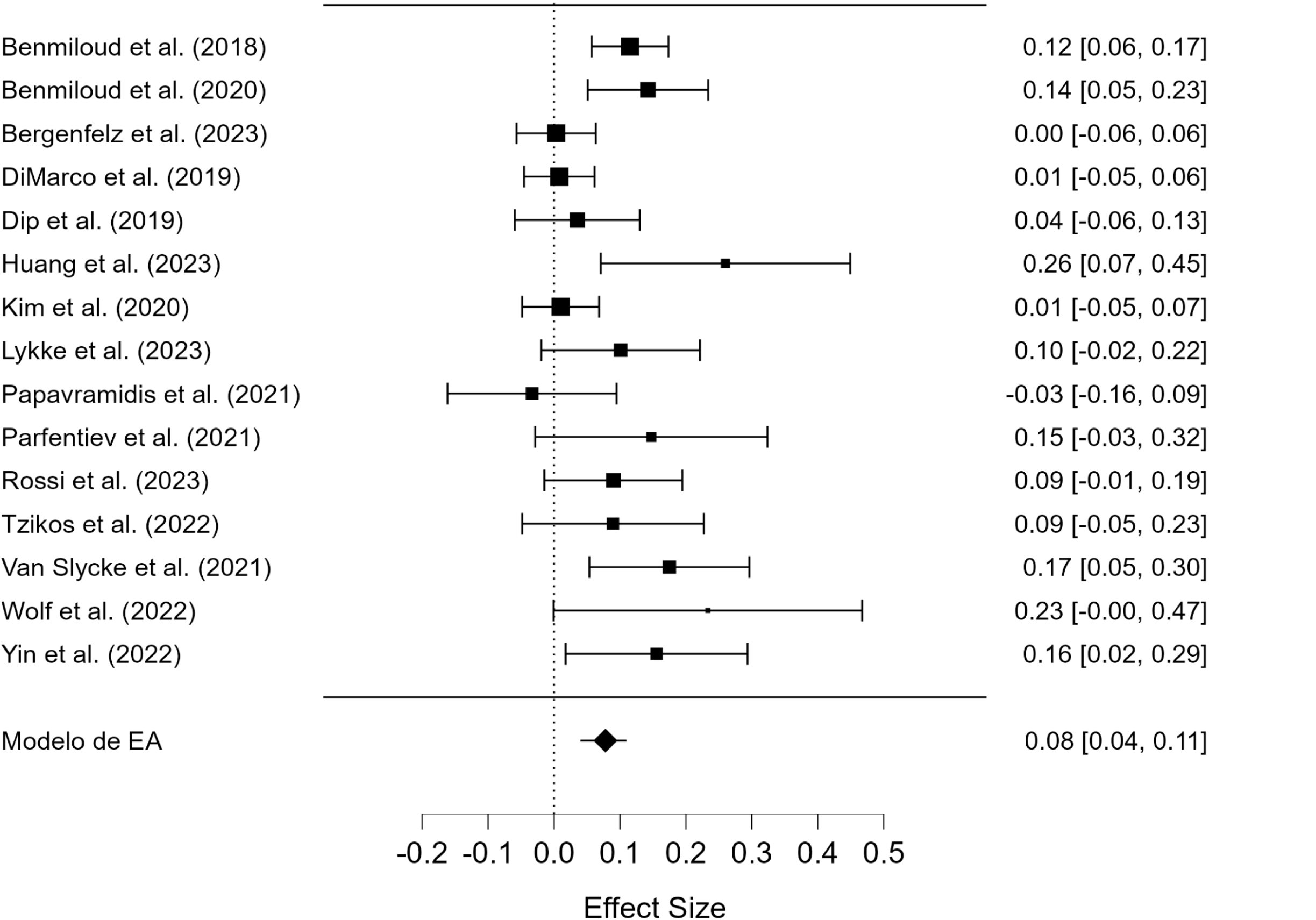
Forest Plot Graph. Hypocalcemia in the sample. Difference in proportions in the NIRAF group versus control.

**Figure 3.**
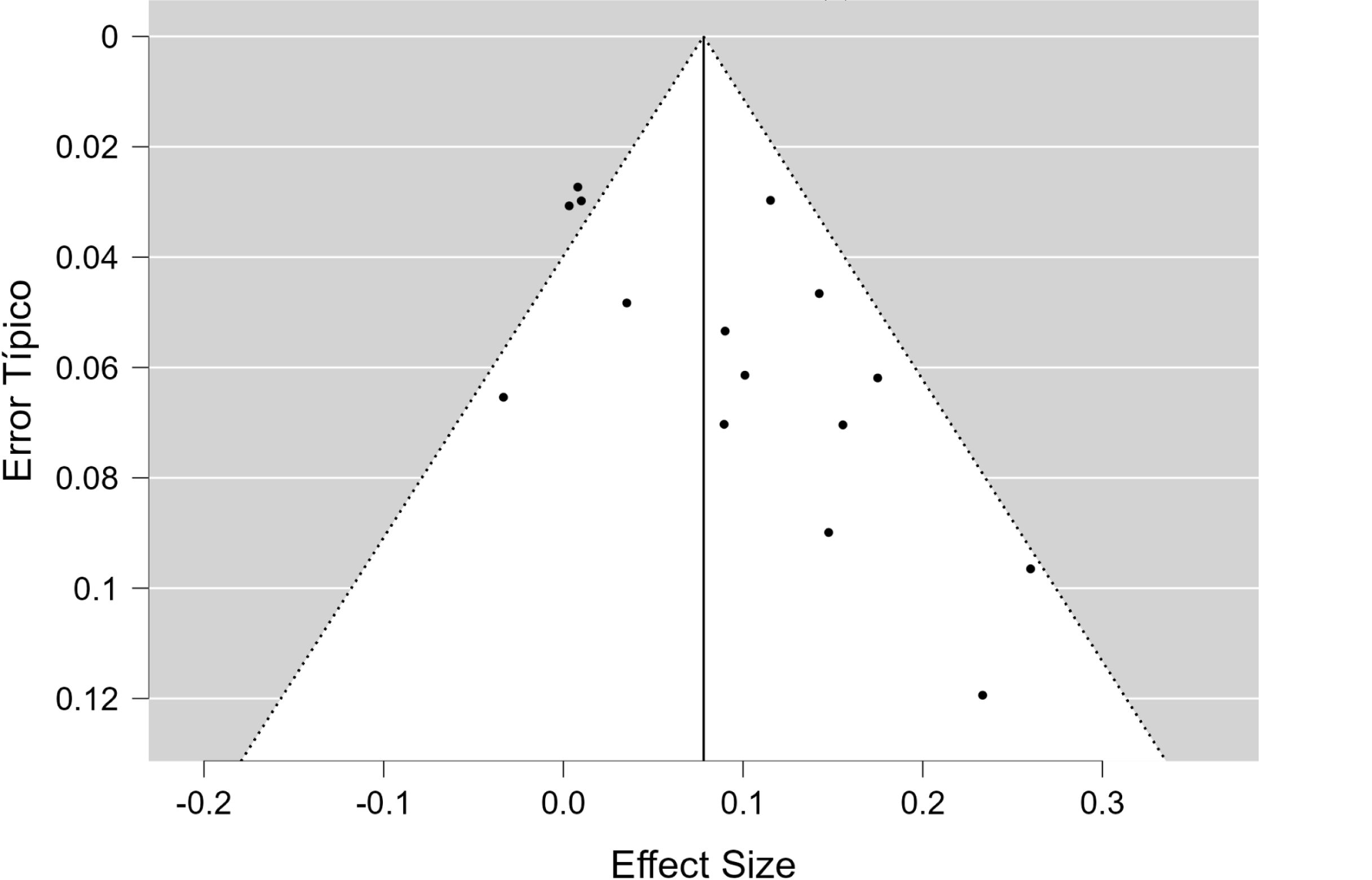
Funnel Plot chart. Hypocalcemia in the sample. Difference in proportions in the NIRAF group versus control.

**Table 7.** Hypocalcemia in the sample. Difference in proportions in the NIRAF group versus control.

|  |  |  |  |  |  |  |
| --- | --- | --- | --- | --- | --- | --- |
| *Fixed and Random Effects* | Q | GI | p |  |  |  |
| Omnibus Contrast of Model Coefficients | 17.09 | 1 | 3.57×10-5 |  |  |  |
| Contrast of the Heterogeneity of Errors | 32.33 | 14 | 3.60×10-3 |  |  |  |
| *Note.* The model was estimated using the Maximum Likelihood method. |  |  |  |  |  |  |
| *Note.* The p-values are approximate. |  |  |  |  |  |  |
| *Coefficients* | Estimate | Typical Error | z | p | 95% Lower CI | 95% Upper CI |
| Intercept | 0.08 | 0.02 | 4.13 | 3.57×10-5 | 0.04 | 0.11 |
| *Adjustment measures* | MV |  |  |  |  |  |
| Log-Verisimilitude | 17.01 |  |  |  |  |  |
| Deviation | 25.65 |  |  |  |  |  |
| AIC | -30.02 |  |  |  |  |  |
| BIC | -28.60 |  |  |  |  |  |
| Aicc | -29.02 |  |  |  |  |  |
| *Estimates of Error Heterogeneity* | Estimate | 95% Lower CI | 95% Upper CI |  |  |  |
| $\tau^2$ | 2.44×10-3 | 4.20×10-4 | 0.01 | | | |
| $\tau$ | 0.05 | 0.02 | 0.11 | | | |
| $I^2$ (%) | 53.13 | 16.35 | 85.93 | | | |
| $H^2$ | 2.13 | 1.20 | 7.11 | | | |
| *Covariances of Parameters* | Intercept |  |  |  |  |  |
|  | 3.56×10-4 |  |  |  |  |  |
| *Rank Correlation Contrast for Asymmetry with Chart in Funnel Plot* | $\tau$ from Kendall | p | | | | |
|  | 0.37 | 0.06 |  |  |  |  |
| *Regression Contrast for Graph Asymmetry in Funnel Plot ("Egger's Probe")* | z | P |  |  |  |  |
|  | 2.93 | 3.35×10-3 |  |  |  |  |

There are significant differences in the proportions of hypocalcemia between the NIRAF and control groups, with moderate to high heterogeneity in errors and possible asymmetry in the data, suggesting variability between the studies included in the analysis and the possible presence of publication bias.

The results of the analysis of transient hypocalcemia in the sample using the difference in proportions between the NIRAF group and the control group are summarized in Table 8, Figure 4 (Forest plot) and Figure 5 (Funnel plot).

**Figure 4.**
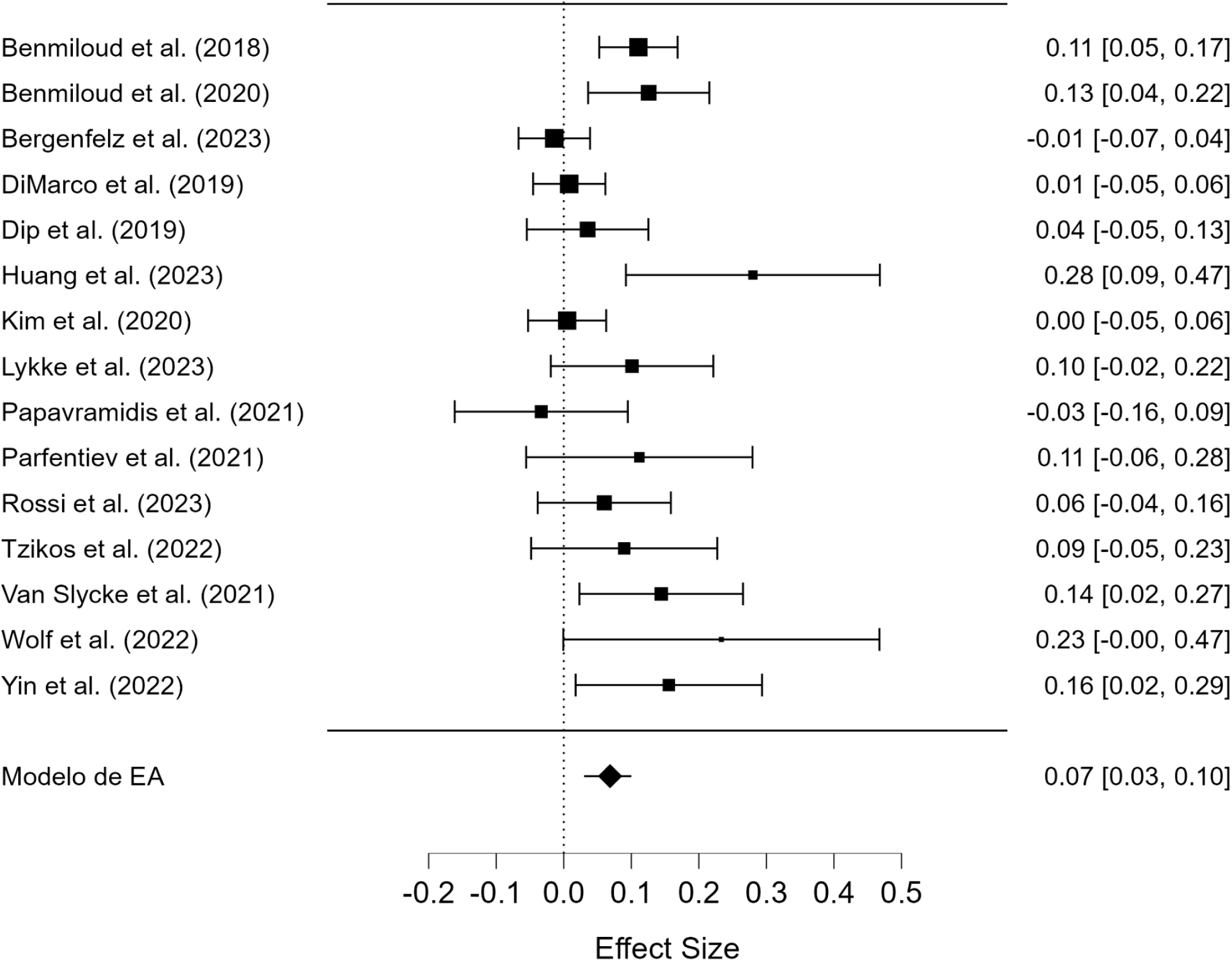
Forest Plot Graph. Transient hypocalcemia in the sample. Difference in proportions in the NIRAF group vs. control.

**Figure 5.**
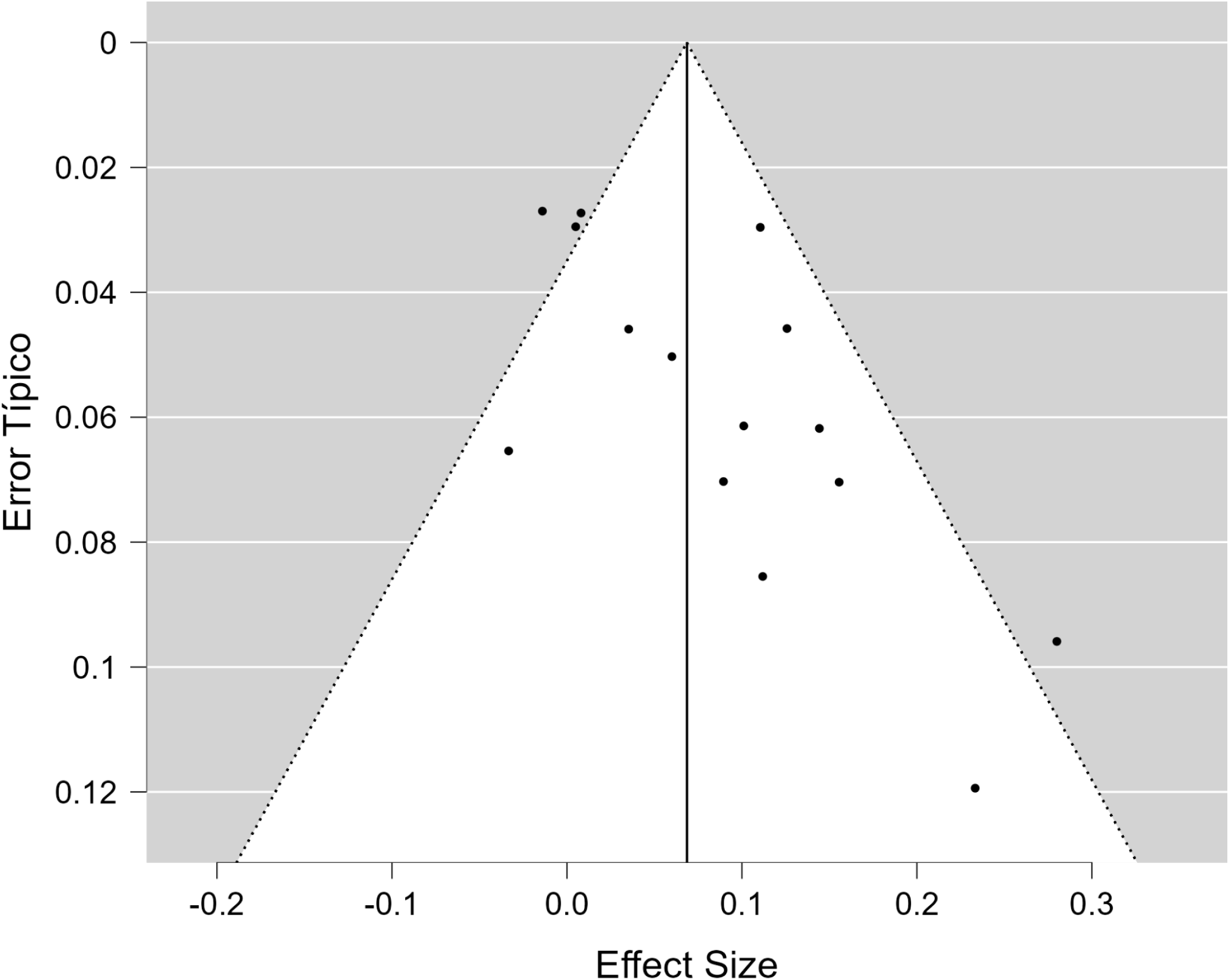
Funnel Plot chart. Transient hypocalcemia in the sample. Difference in proportions in the NIRAF group versus control.

**Table 8.** Transient hypocalcemia in the sample. Difference in proportions in the NIRAF group vs. control.

|  |  |  |  |  |  |  |
| --- | --- | --- | --- | --- | --- | --- |
| *Fixed and Random Effects* | Q | GI | p |  |  |  |
| Omnibus Contrast of Model Coefficients | 13.82 | 1 | 2.01×10-4 |  |  |  |
| Contrast of the Heterogeneity of Errors | 33.21 | 14 | 2.69×10-3 |  |  |  |
| *Note.* The model was estimated using the Maximum Likelihood method. |  |  |  |  |  |  |
| *Note.* The p-values are approximate. |  |  |  |  |  |  |
| *Coefficients* | Estimate | Typical Error | z | p | 95% Lower CI | 95% Upper CI |
| Intercept | 0.07 | 0.02 | 3.72 | 2.01×10-4 | 0.03 | 0.10 |
| *Note.* Wald's contrast. |  |  |  |  |  |  |
| *Adjustment measures* | MV |  |  |  |  |  |
| Log-Verisimilitude | 17.10 |  |  |  |  |  |
| Deviation | 26.13 |  |  |  |  |  |
| AIC | -30.20 |  |  |  |  |  |
| BIC | -28.78 |  |  |  |  |  |
| Aicc | -29.20 |  |  |  |  |  |
| *Estimates of Error Heterogeneity* | Estimate | 95% Lower CI | 95% Upper CI |  |  |  |
| $\tau^2$ | 2.32×10-3 | 4.39×10-4 | 0.01 | | | |
| $\tau$ | 0.05 | 0.02 | 0.12 | | | |
| I<sup>2</sup> (%) | 53.41 | 17.82 | 87.08 |  |  |  |
| H<sup>2</sup> | 2.15 | 1.22 | 7.74 |  |  |  |
| *Covariances of Parameters* | Intercept |  |  |  |  |  |
|  | 3.40×10-4 |  |  |  |  |  |
| *Rank Correlation Contrast for Asymmetry with Chart in Funnel Plot* | $\tau$ from Kendall | p | | | | |
|  | 0.41 | 0.04 |  |  |  |  |
| *Regression Contrast for Graph Asymmetry in Funnel Plot ("Egger's Probe")* | z | P |  |  |  |  |
|  | 3.11 | 1.87×10-3 |  |  |  |  |

The results indicate that there are significant differences in the proportions of transient hypocalcemia between the NIRAF and control groups, with moderate to high heterogeneity in errors and possible asymmetry in the data, suggesting variability between the studies included in the analysis and the possible presence of publication bias.

The results of the analysis of permanent hypocalcemia in the sample using the difference in proportions between the NIRAF group and the control group are summarized in Table 9, Figure 6 (Forest plot) and Figure 7 (Funnel plot).

**Figure 6.**
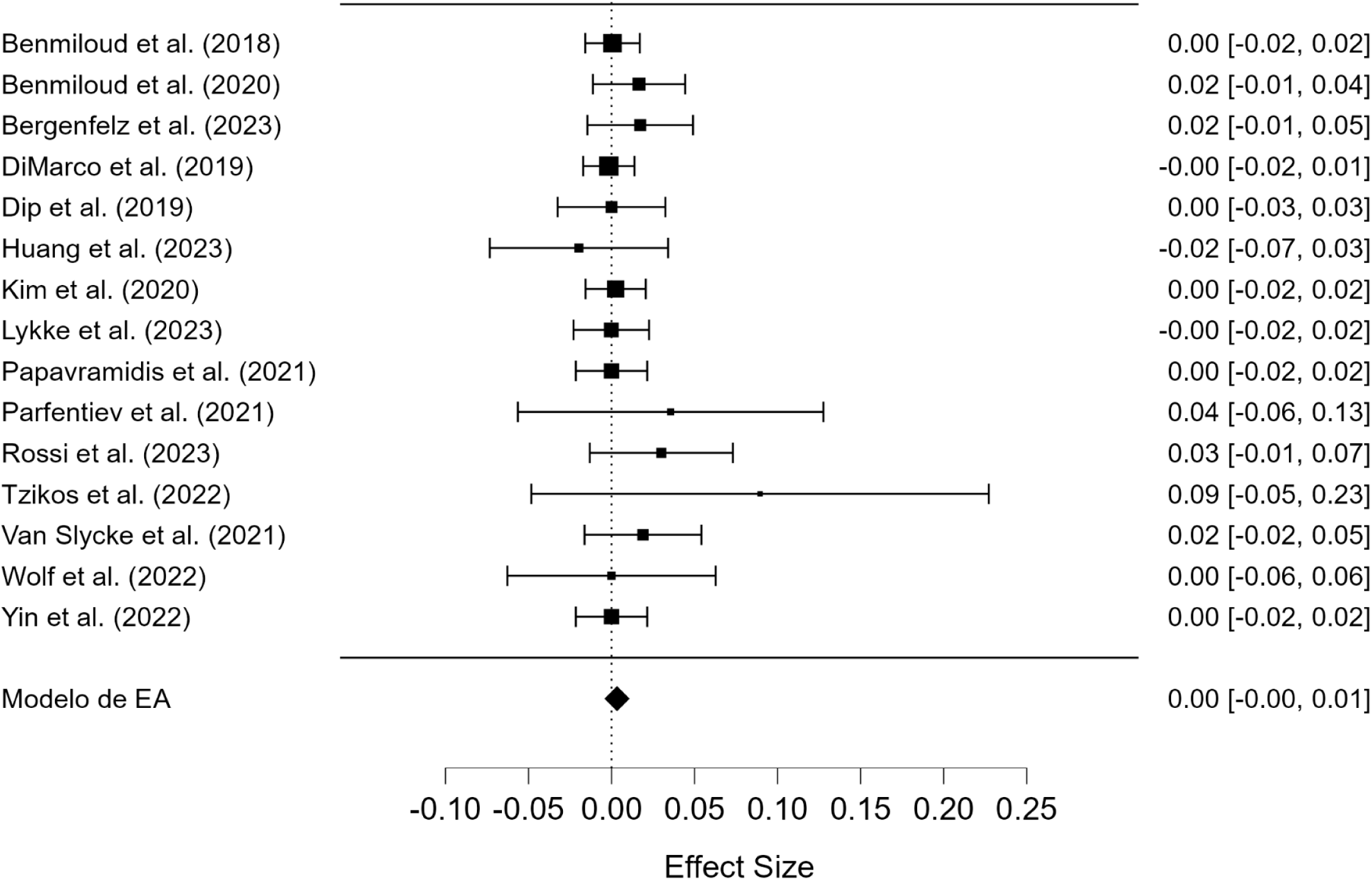
Forest Plot Graph. Permanent hypocalcemia in the specimen. Difference in proportions in the NIRAF group versus control.

**Figure 7.**
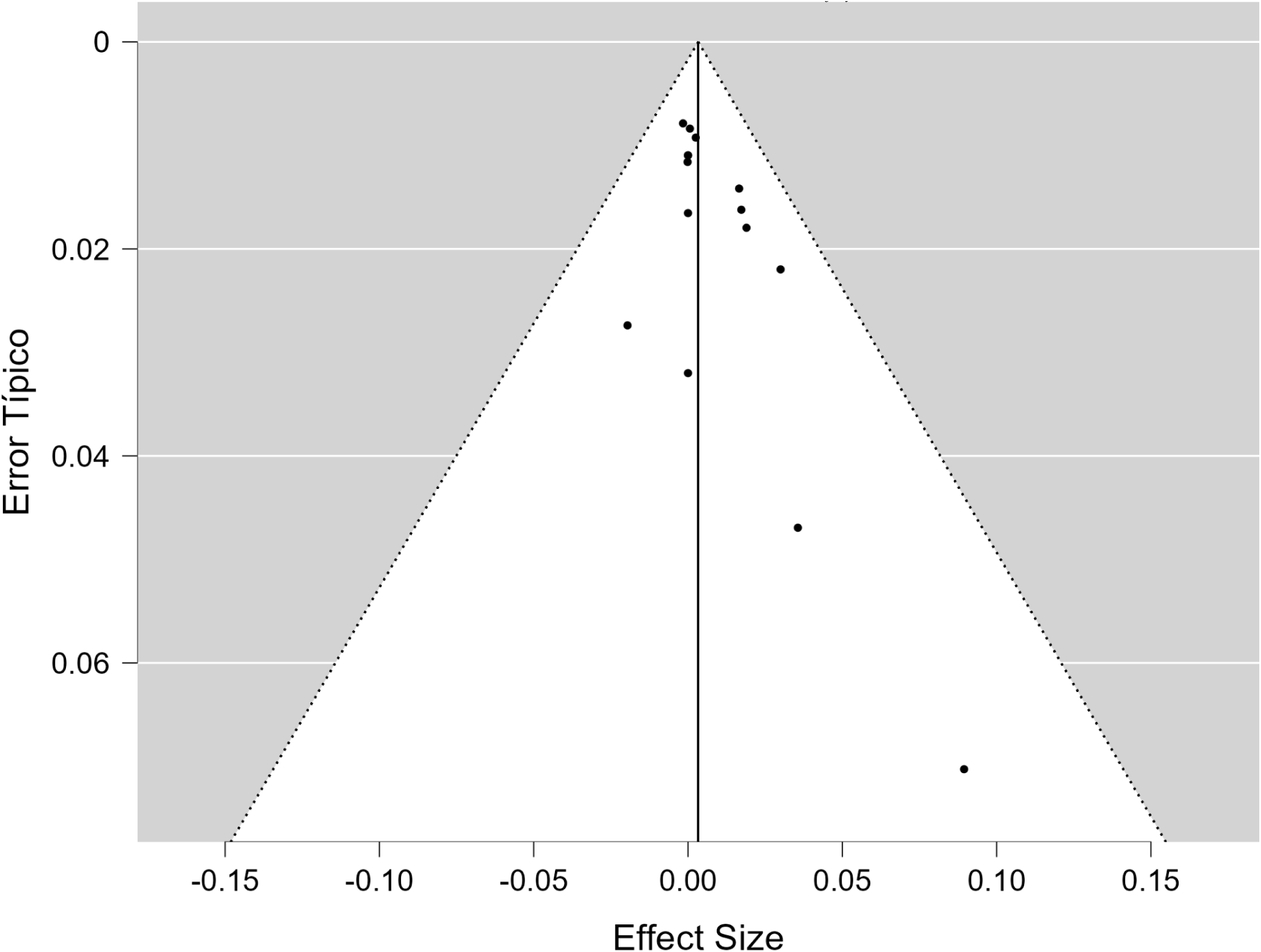
Funnel Plot chart. Permanent hypocalcemia in the specimen. Difference in proportions in the NIRAF group versus control.

**Table 9.**
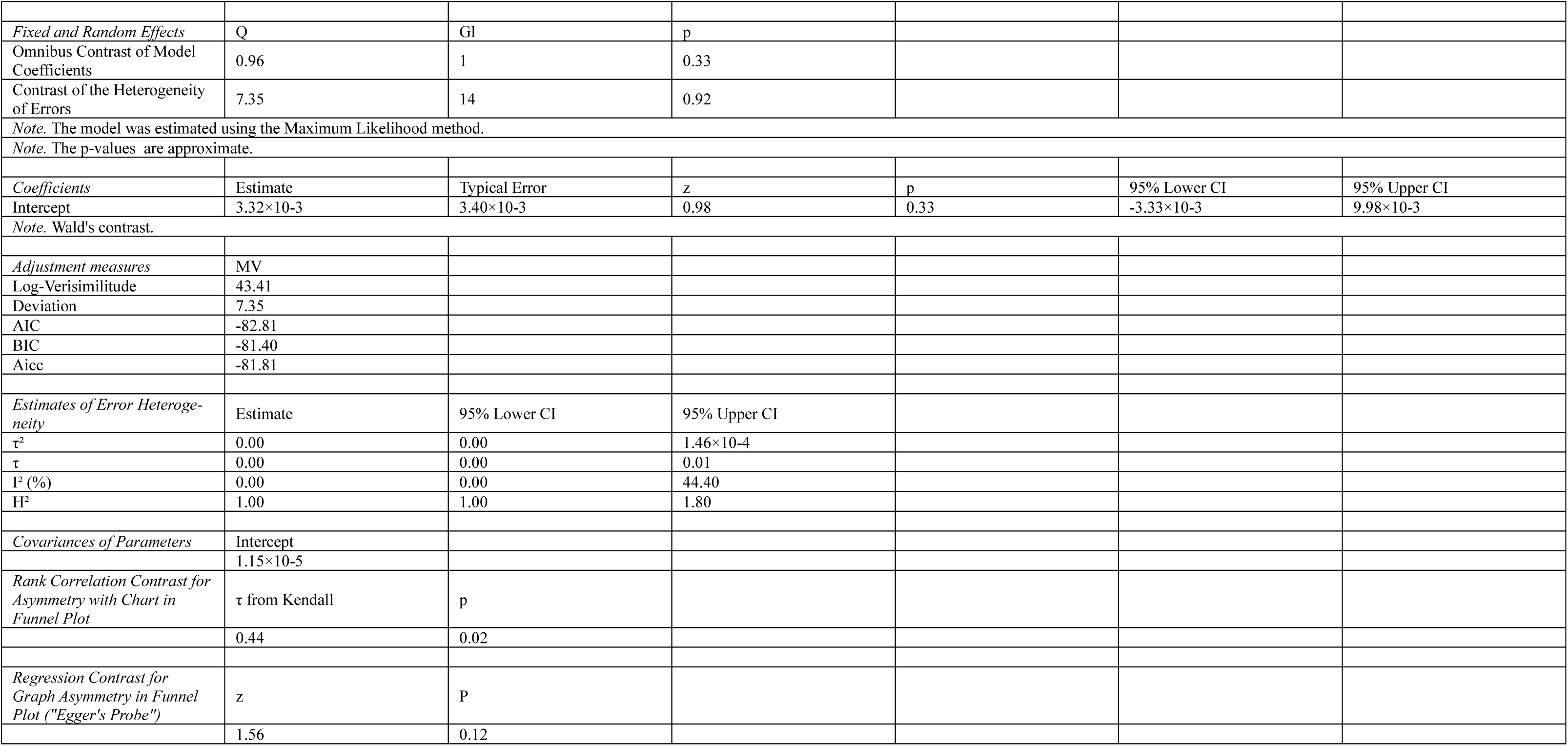
Permanent hypocalcemia in the specimen. Difference in proportions in the NIRAF group vs. control.

The results indicate that there are no significant differences in the proportions of permanent hypocalcemia between the NIRAF and control groups, with homogeneity in errors and no significant evidence of publication bias.

Research biases

Figures 8 and 9 show the assessment of the risk of research bias with the ROBINS-I scale. Overall, research bias is mild-moderate.

**Figure 8.**
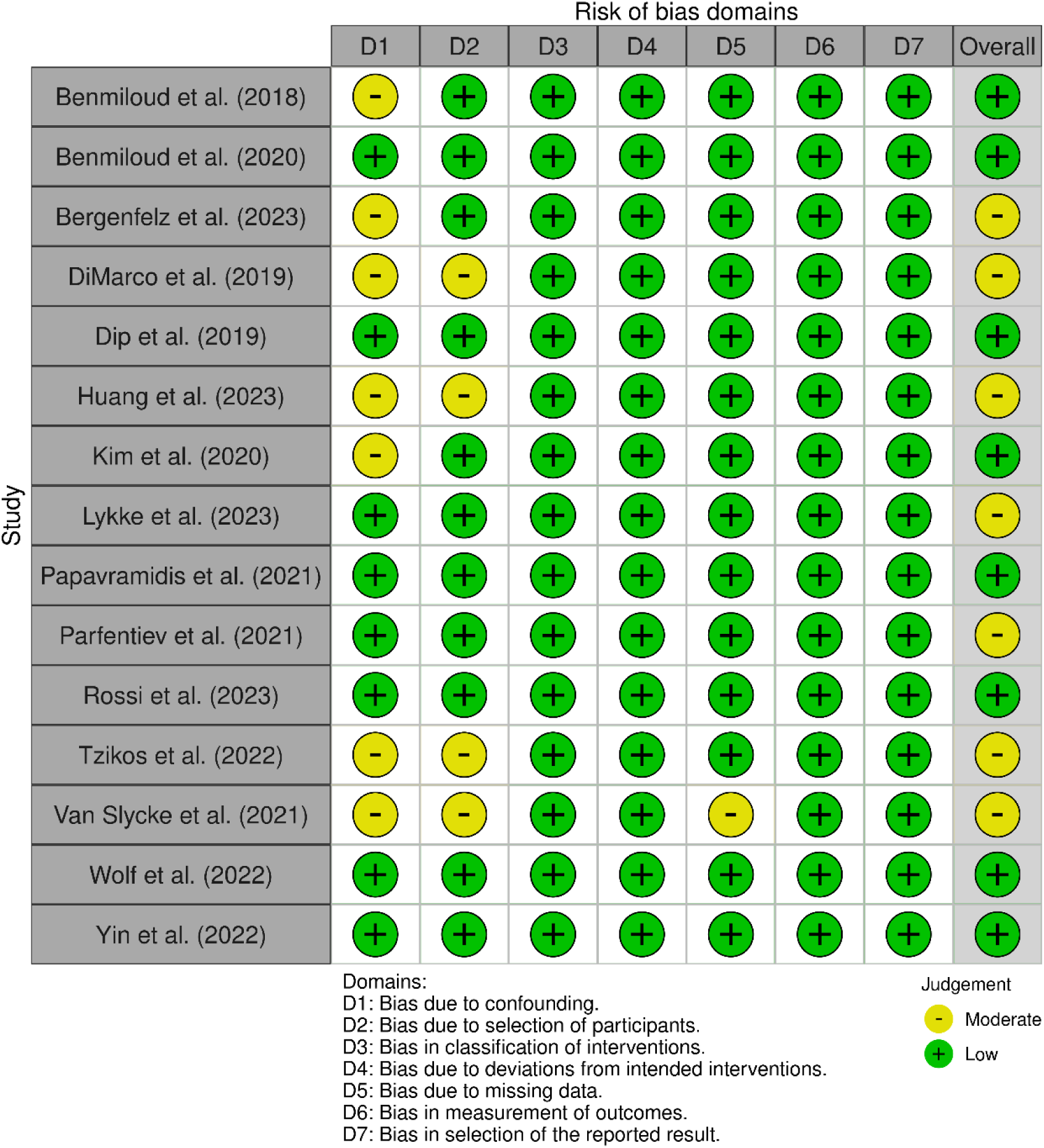
Evaluation of publication biases with the ROBINS-I scale of the dimensions for each of the selected articles.

**Figure 9.**
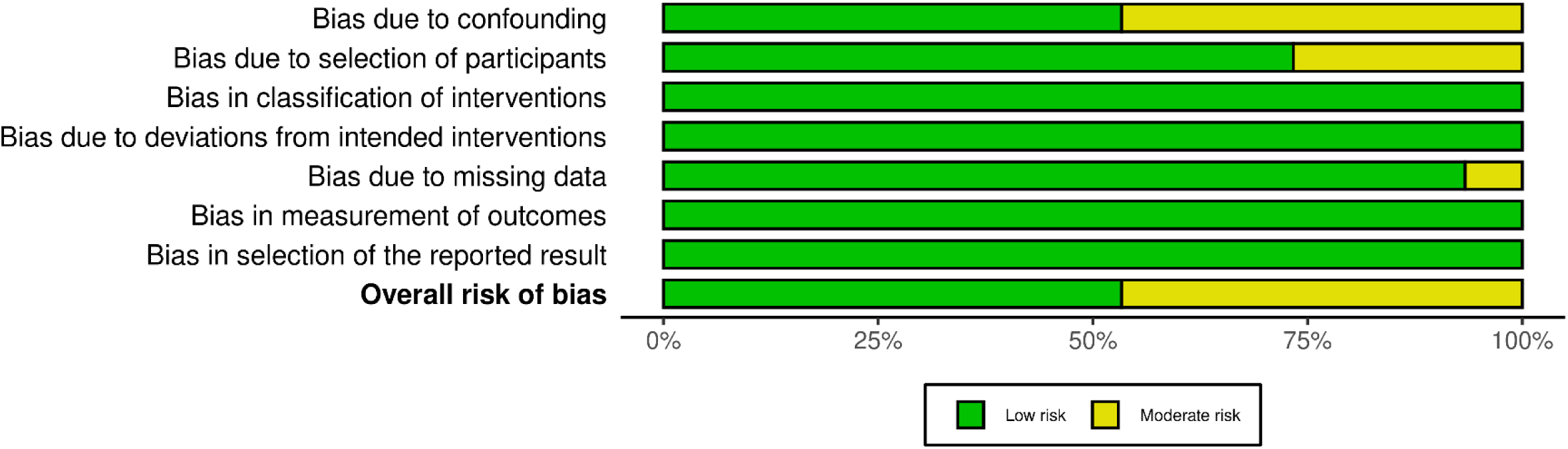
Evaluation of publication biases with the ROBINS-I scale of the dimensions for all the selected articles.

Level of evidence and degree of recommendation (GRADE)

Table 10 summarizes the evaluation of each of the articles that were included in the meta-analyses. The studies by Benmiloud et al. (2020), Dip et al. (2019), and Rossi et al. (2023) are notable for their high methodological quality and their results suggest that NIRAF can significantly reduce the risk of postoperative hypocalcemia, increase the identification of the parathyroid glands, and reduce the rates of autotransplantation and inadvertent resection of these glands [10,14,20].

**Table 10.** Evaluation of the level of evidence and degree of recommendation with the GRADE system.

| Article | Study Design | Risk of Bias | Consistency | Direct Evidence | Precision | Level of Evidence (GRADE) | Strength of Recommendation (GRADE) |
| --- | --- | --- | --- | --- | --- | --- | --- |
| Benmiloud et al., 2018 | Controlled before and after study | Moderate (possible Hawthorne bias) | Moderate (consistent results) | Direct | Moderate | Casualty | Weak |
| Benmiloud et al., 2020 | Randomized Clinical Trial | Low | Discharge (multicenter) | Direct | Loud | Loud | Strong |
| Bergenfelz et al., 2023 | Randomized Clinical Trial | Moderate (inter-site variability) | Casualty | Direct | Moderate | Moderate | Weak |
| DiMarco et al., 2019 | Retrospective study | High | Casualty | Innuendo | Casualty | Very Low | Weak |
| Dip et al., 2019 | Randomized Clinical Trial | Low | Loud | Direct | Moderate | Loud | Strong |
| Huang et al., 2023 | Narrative Review | N/A | N/A | Innuendo | N/A | N/A | Weak |
| Kim et al., 2020 | Before and after study | Moderate | Casualty | Direct | Casualty | Casualty | Weak |
| Lykke et al., 2023 | Randomized Clinical Trial | Moderate | Moderate | Direct | Moderate | Moderate | Weak |
| Papavramidis et al., 2021 | Randomized Clinical Trial | Moderate | Moderate | Direct | Moderate | Moderate | Weak |
| Parfentiev et al., 2021 | Comparative prospective study | Moderate | Casualty | Direct | Casualty | Casualty | Weak |
| Rossi et al., 2023 | Randomized Clinical Trial | Low | Loud | Direct | Loud | Loud | Strong |
| Van Slycke et al., 2021 | Retrospective study | High | Casualty | Direct | Casualty | Very Low | Weak |
| Wolf et al., 2022 | Randomized Clinical Trial | Moderate | Casualty | Direct | Moderate | Moderate | Weak |
| Yin et al., 2022 | Comparative prospective study | Moderate | Moderate | Direct | Moderate | Moderate | Weak |

Study Design: Classification of the type of study (randomized clinical trial, observational study, etc.).
Risk of Bias: Assessment of the risk of bias inherent in the study design (high, moderate, low).
Consistency: Evaluation of the consistency of results with other studies (high, moderate, low).
Direct Evidence: Indicates whether the evidence is direct to the question posed (direct, indirect)
Accuracy: Evaluation of the accuracy of results (high, moderate, low).
Level of Evidence (GRADE): Classification of the overall quality of the evidence (high, moderate, low, very low).
Strength of Recommendation (GRADE): Strength of the evidence-based recommendation (strong, weak).

The current evidence-based global recommendation is that the use of NIRAF can be considered as a useful tool in total thyroidectomy to improve the identification and preservation of the parathyroid glands, although the strength of the recommendation may be weak in favour due to heterogeneity, risk of bias, inconsistent results, and studies with very low evidence.

## Discussion

### Regarding meta-analyses [7–9] (Tables 1 and 2)

The interpretation of the results obtained in meta-analyses should be interpreted with caution due to heterogeneity in some studies when combining RCTs with investigations with other methodologies.

Meta-analyses by Rao et al. and Safia et al. collect exclusively in RCTs [7,8].

Safia et al. found that NIRAF is associated with higher postoperative PTH levels, higher PG identification rate, higher postoperative serum calcium levels, and a reduction in inadvertent removal of parathyroid glands [8].

The definition of hypocalcemia varies from study to study, and there is variability in the determination and criteria for permanent hypoparathyroidism.

Meta-analyses by Rao et al. and Weng et al. [7,9] showed no apparent publication bias according to the *Funnel Plots*. Safia et al. did not assess publication bias due to the small number of studies [8].

The devices used and their application vary from one study to another, which may introduce variability in the results as there is no standardization of the NIRAF technique.

The studies included in the meta-analyses are based on techniques performed by experienced surgeons, which may limit the generalization of the technique to less experienced surgeons.

Meta-analyses do not directly assess the cost-effectiveness of the NIRAF technique, which is a major limitation to its widespread implementation. More research is required to determine whether the clinical benefits justify the additional costs.

### Regarding meta-meta-analysis [10–24] (Tables 5 to 9; Figures 2 to 7)

There is a significant association between the presentation of transient (and global) postoperative hypocalcemia after total thyroidectomy and the use or not of the NIRAF system (Table 5).

The prevalence of transient hypocalcemia is significantly lower in the NIRAF groups (Tables 7 and 8, Figures 2 to 5).

The prevalence of permanent hypocalcemia, although also lower, does not have a statistically significant difference (Table 9, Figures 6 and 7).

There is publication bias when assessing the prevalence of global hypocalcemia for the sample (Figure 3) or transient hypocalcemia (Figure 5). There is no publication bias in assessing the prevalence of permanent hypocalcemia (Figure 7)

Potential research biases were assessed with ROBINS-I which, although useful for comparison, does not replace the specific assessment of risk of bias that would be done with RoB2 for RCTs or NOS for observational studies.

The interpretation of the results, among others, may be influenced by several factors:

- The criterion used to define hypocalcemia or hypoparathyroidism.
- The NIRAF device used.
- Other factors that can influence damage to the parathyroid glands.

### Criteria for defining hypocalcemia

Postoperative hypocalcemia is assumed to be caused by hypoparathyroidism after total thyroidectomy caused by manipulation of the parathyroid glands, their devascularization, or their inadvertent resection [10,11,18,19,23,24]. Preservation of the parathyroid glands and their vascularization is crucial to avoid postoperative hypocalcemia [16,20] and it is assumed that a single intact gland is sufficient to maintain normal levels of calcium and PTH in the blood, and that preservation of as many glands as possible minimizes the incidence of postoperative hypoparathyroidism [19].

The criteria for defining hypocalcaemia are not common to all studies, making it difficult to compare the results. There are discrepancies in the measurement of calcemia (some use a threshold value measuring corrected calcium and others the total), others include the presence of symptoms in the definition, and there is variability in when calcium levels are measured after surgery. There is also no clear reference to the criteria followed in the measurement of postoperative PTH.

Benmiloud et al., 2018 consider hypoparathyroidism in patients with a corrected calcium level <8.0 mg/dL (approximately <2 mmol/L) on day 1 or 2 postoperatively [11] [16], considering severe hypocalcemia a serum calcium level less than 7.6 mg/dL [10,14]. Lykke et al., 2023 define hypoparathyroidism as the presence of hypocalcemia (ion-Ca2+ < 1.15 mmol/L) with low PTH levels and the need for continuous treatment with active vitamin D and consider it permanent if it lasts 6 months or more, or if continuous treatment with calcium and active vitamin D is needed [17].

Tzikos et al., 2022 defines hypocalcemia as a serum calcium level below 2 mmol/l (8.0 mg/dL.) and PTH <1 pmol/l (9.43 pg/mL) in patients requiring calcium and vitamin D supplementation for more than two weeks [21].

These definitions show variations in the exact criteria used to define hypoparathyroidism, but all focus on the presence of hypocalcemia and/or low parathyroid hormone levels, as well as the need for calcium and vitamin D supplementation [16–18].

### NIRAF Devices

The devices used are different, the most commonly used being Fluobeam®, followed by Elevision® and the Karl Storz system (see Table 3). The lack of information in some studies on the device used may make it difficult to compare the results and evaluate the effectiveness of different NIRAF technologies.

All systems rely on the emission of the parathyroid glands from near-infrared (NIR) autofluorescence when exposed to NIR light, allowing the glands to be identified during thyroid surgery, helping to prevent accidental resection [11,12,14,18,22–24]. In some studies, it is associated with indocyanine green angiography (ICG) that when exposed to NIR light, the ICG fluoresces, allowing blood vessels to be visualized. In thyroid surgery, ICG can be used to evaluate the perfusion of the parathyroid glands to identify viable glands [19,20,24].

The combination of NIRAF and ICG can provide complementary information on the identification and viability of the parathyroid glands and increase their preservation [24].

### Other factors that can influence parathyroid gland damage

The authors identify factors that may interfere with the identification of parathyroid glands with NIRAF, such as limited penetration of light if the glands are covered by adipose tissue or blood [20]. In addition, with some NIRAF systems it is necessary to interrupt the intervention to use the device, which may not minimize damage to the parathyroid glands, and interpretation of indocyanine green angiography may be subjective [19].

However, autofluorescence during surgery helps identify and preserve the parathyroid glands, thus reducing the risk of hypocalcemia with all devices [10,20].

The hypofunction of the parathyroid glands, which results in hypocalcemia, is influenced by other factors independent of their identification and conservation, such as the preservation of their vascularization [10] and the skill and experience of the surgeon [15]. Difficulty during dissection (extent of surgery, complex anatomy) may increase non-identification and inadvertent removal of the parathyroid glands [16], as well as anatomical variations with ectopic or intrathyroid glands, intraoperative bleeding, or the presence of fat that may make detection with NIRAF difficult in systems with low tissue penetration [20].

Preoperative vitamin D deficiency may contribute to postoperative hypocalcemia, although this does not indicate actual hypoparathyroidism [17].

In summary, damage to the parathyroid glands is multifactorial and depends on the interaction of factors related to the surgical technique, the patient’s anatomy, and the use of visualization tools [22] that facilitate the preservation of parathyroid gland vascularization, such as indocyanine green angiography [24].

### Level of evidence and degree of recommendation (GRADE)

Some of the randomized clinical trials (RCTs) analyzed have shown promising results, but the variability between centers and series involving the same expert surgeons [11,14,23] require new multicenter RCTs with larger numbers of patients to confirm the efficacy of autofluorescence in different settings and with different surgeons [12]. These studies must be sufficiently powered to detect significant differences in permanent hypocalcemia and not just transient hypocalcemia [17]. Studies should consider differences in inclusion and exclusion criteria, such as preoperative vitamin D levels, and look for uniformity in postoperative calcium and PTH sampling.

In summary, in total thyroidectomy NIRAF may be useful in identifying and preserving the parathyroid glands, but more trials and different literature research approaches are needed to confirm these findings and establish autofluorescence as a standard.

### Future lines of research

Various lines of research are suggested to improve the accuracy of the NIRAF, deduced from the studies analyzed, such as:

- Optimization of autofluorescence technology.
- Investigate the combination of NIRAF with other techniques, such as indocyanine green angiography (ICG) to assess vascularisation or other imaging techniques such as the use of laser mottled contrast.
- Long-term impact assessment with longitudinal follow-ups to determine the incidence of permanent hypoparathyroidism with a clear and standardized definition of permanent hypoparathyroidism to facilitate comparisons between investigations.
- Analysis of the learning curve and usefulness in surgeons with different levels of experience.

### Limitations of the study

- This is a meta-meta-analysis that includes only the studies included in three initially selected metaanalyses [7–9].
- Study design and selection bias due to the inclusion of studies with different methodologies (RCTs, retrospective studies, non-randomised studies, studies involving a single surgeon or a single centre) [11,13,14,16,19,22,23].
- Insufficient sample sizes [16,21–23].
- Heterogeneity and variability between studies [12,16,17].
- Limitations in the handling and interpretation of the NIRAF technique [22] such as false negatives, the presence of fatty tissue or difficulty in observing marginal areas.
- Lack of long-term follow-up to assess the actual rate of permanent hypoparathyroidism [16,23].
- Possible Hawthorne effect (unconscious change in surgeon’s attitude to participating in a study) [11,22,24].

### Conclusions

Near-infrared light autofluorescence (NIRAF) improves identification of the parathyroid glands during total thyroidectomy.

NIRAF is useful for identifying and preserving the parathyroid glands during total thyroidectomy. The presentation of postoperative hypocalcemia is associated with the use or non-use of NIRAF.

In the group of patients in whom NIRAF was used, the prevalence of global and transient postoperative hypocalcemia is significantly lower.

The current evidence-based global recommendation is that the use of NIRAF can be considered as a useful tool in total thyroidectomy to improve the identification and preservation of the parathyroid glands, although the strength of the recommendation may be weak in favour due to heterogeneity, risk of bias, inconsistent results, and studies with very low evidence.

## Data Availability

All data produced in the present work are contained in the manuscript

## Notes

### Competing Interest Statement

The authors have declared no competing interest.

### Funding Statement

This study did not receive any funding

